# Randomized Clinical Trial of ICECaP (Individualized Coordination and Empowerment for Care Partners of Persons with Dementia): Primary Mental Health and Burden Outcomes

**DOI:** 10.1101/2024.08.15.24312041

**Authors:** Virginia T. Gallagher, Anna Arp, Ryan Thompson, Agustina Rossetti, James Patrie, Shannon E Reilly, Carol A. Manning

## Abstract

Efficacy of the Individualized Coordination and Empowerment for Care Partners of Persons with Dementia (ICECaP), an intervention that involves one-on-one individualized support from a dementia care coordinator for a dementia care partner, compared to an active control group. At least once monthly contact is made from a dementia care coordinator to the dementia care partner by telephone, video conferencing, email, or in-person support at clinical visits for the person with dementia. In this pilot randomized unblinded control trial of ICECaP, n=61 (n=90 randomized) care partners completed 12-months of the ICECaP intervention and n=69 (n=92 randomized) care partners received routine clinical support (controls) in an outpatient memory care clinic at an academic medical center, from which the participants were recruited (ClinicalTrials.gov: NCT04495686, funded by Department of Defense and Virginia Department for Aging and Rehabilitative Services). Early termination endpoints (death and higher level of care) and trial drop out were comparable across groups. Primary efficacy outcomes were evaluated by comparing changes in care partner mental health, burden, and quality of life from baseline to 12-months between ICECaP and controls. Linear-mixed ANCOVA revealed no significant group differences in longitudinal changes on measures of caregiving burden, care partner depression, anxiety, quality of life, or reactions to the behavioral symptoms of the person with dementia. Hypothesized reasons for lack of initial efficacy on primary 12-month outcomes are discussed.

## Introduction

There are over 11 million family members and friends providing unpaid care for persons with dementia in the United States. Informal dementia caregiving is valued at $350 billion and 18.4 billion hours of care per year nationally.^1^ Although many dementia care partners experience meaning and fulfillment in the context of their caregiving role, caring for a person with dementia is associated with increased levels of psychosocial stress, depression, and emotional burden.^2–9^

Many programs and interventions have been developed to support dementia care partners and improve their mental health, quality of life, and caregiving readiness. Among them, collaborative care coordination has emerged as a promising, individualized intervention to help care partners and their care recipients with dementia navigate complex health systems, financial/insurance systems, and community resources.

Additionally, care partners are provided with social and emotional support.^10–13^ A team at the University of Virginia, along with its partners, developed an intervention for dementia care partners called ICECaP: Individualized Coordination and Empowerment for Care Partners of Persons with Dementia to support dementia care partners. ICECaP involves individualized elements of care coordination, supportive counseling, psychoeducation, and skills training and is delivered in a hybrid setting – combining an optional initial home visit; ongoing, at least-monthly telehealth interactions via phone, email, and HIPAA-compliant video calls; and accompaniment to clinic visits for the person with dementia.

In this article, we report preliminary, primary efficacy outcomes for a pilot, randomized unblinded control trial (RCT) of ICECaP. Please see Gallagher et al. 2024 for program details, the ICECaP protocol, and analytical plan.^14^ We evaluate Aim 1 of the ICECaP RCT: determine whether ICECaP improves care partner burden, symptoms of depression, reaction to the behavioral symptoms of dementia, and quality of life. We hypothesized that after controlling for baseline characteristics, including level of care-recipient functional dependence, care partners in the ICECaP group would significantly improve from baseline to 12 months (post-intervention) on mental health and quality of life measures, whereas controls would not improve.

## Methods

This study was approved by the University of Virginia Institution Research Board for Health Services Research and registered with ClinicalTrials.gov (NCT04495686). All care partners underwent a written informed consent process one-on-one with a trained clinical research coordinator (CRC) prior to initiating study procedures. Consent was documented with the signature of the participant and the CRC. All participants were provided with a signed copy of the consent document.

### Recruitment

As reported in the published protocol^14^ and the feasibility and acceptability data for the ICECaP clinical trial (Thompson et al., currently under review), care partners were recruited from March 1, 2021 to September 30, 2022 from the University of Virginia’s multidisciplinary Memory and Aging Care Clinic (MACC) when accompanying a patient with dementia to a clinical appointment. Care partners were required to be aged ≥18 years, possess basic spoken and written English skills, have home-based internet access, and self-identify as the primary care partner for a patient diagnosed with mild to moderate dementia living in the community (e.g., not living in a continuing care facility). During the course of the trial, eligibility criteria was expanded to include patient populations being overlooked for recruitment. Changes made included lowering age requirements, and including multiple etiologies of dementia. Using a random permuted block randomization scheme generated by the study statistician and implemented by the clinical research coordinator at enrollment, care partners were randomly assigned to 12-months of ICECaP or 12-months of routine clinical care (controls).

### Target sample size

Based on an a priori power analysis, the target sample size was n=140 CPs, (50% ICECaP and 50% control) to achieve at least 0.80 power for detecting a small-to-medium Cohen’s *d* effect size (*d* = 0.3 to 0.5) when comparing baseline to 12-month mean change in the care partner’s depression score (Center for Epidemiological Studies Depression Scale–Revised) and when comparing the baseline to 12-month mean change the care partner’s burden score (Zarit Burden Interview) between ICECaP and control groups at a significance level of α = .05.

### Sample and Attrition

As described in Thompson et al. (under review), of the n=169 (control n=87, ICECap n=82) care partners recruited into the RCT who completed baseline assessments (out of 182 participants randomized), 23.08% were withdrawn from the final sample because within 12-months, the person with dementia died (n=9), moved to a higher level of care (n=8), or moved out of state (n=1); or the care partner chose to withdraw (n=4), was lost to follow-up despite two attempts to contact (n=13), or did not complete 12-month assessments (n=4). See Figure 1. The final sample of care partners included n=69 controls and n=61 ICECaP who completed baseline and 12-month assessments.

**Figure 1.**
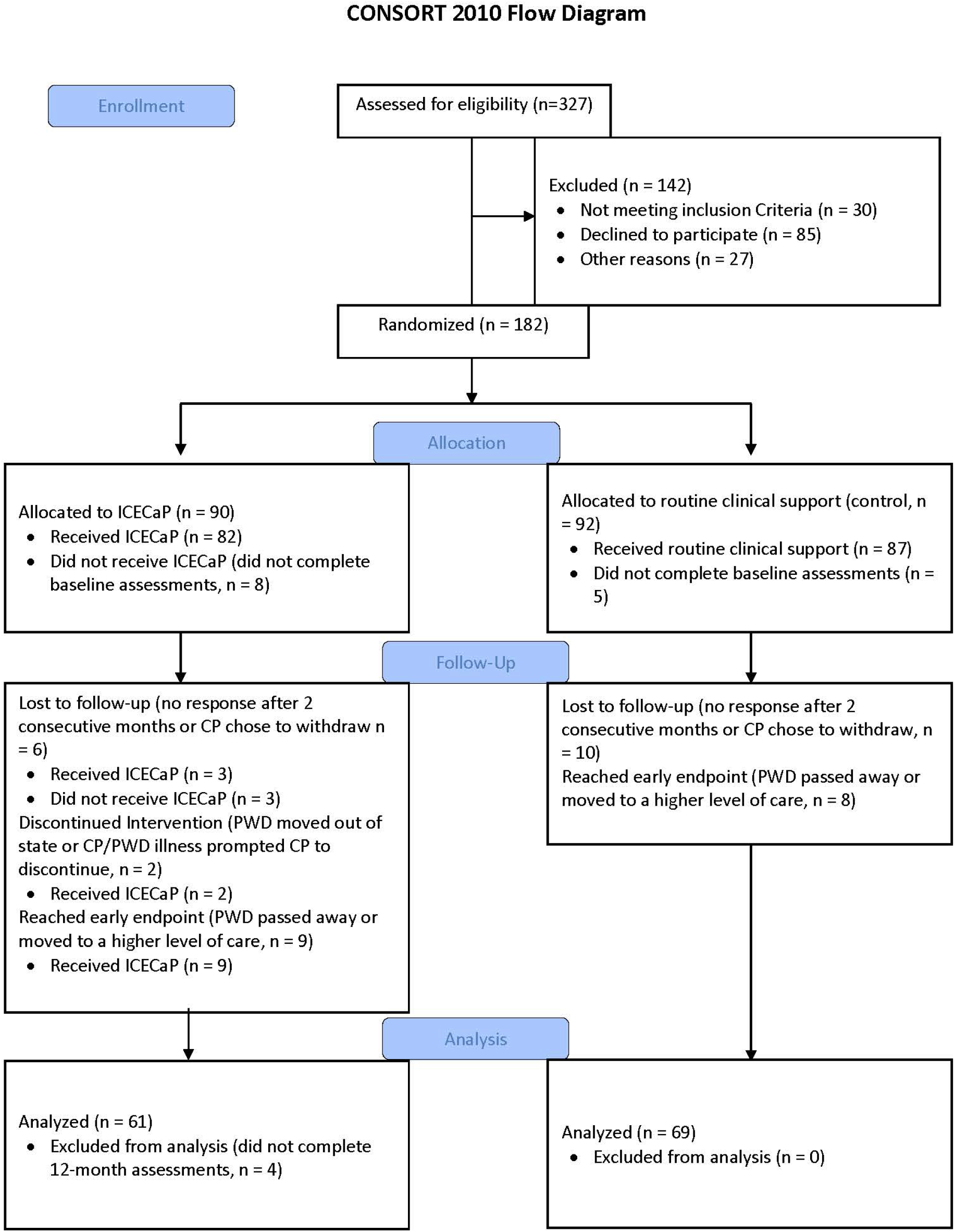
Sample Consort Flow Diagram.

### Procedures and Intervention

Please see Gallagher et al., 2024 for details. Questionnaires listed in Table 1 were completed online by both ICECaP and control groups at baseline and 12-months after baseline. Additional questionnaires for sample characterization included the Katz Index of Independence in ADLs (Katz Basic ADLs),^15^ the Lawton-Brody Instrumental ADL Scale (Lawton-Brody Complex ADLs),^16^ and the Neuropsychiatric Inventory Questionnaire, all completed by the care partner about the person with dementia.^17^ All measures were collected and stored using REDCap^18^, hosted by the University of Virginia, and were monitored by the clinical research coordinator for missing data.

**Table 1.**
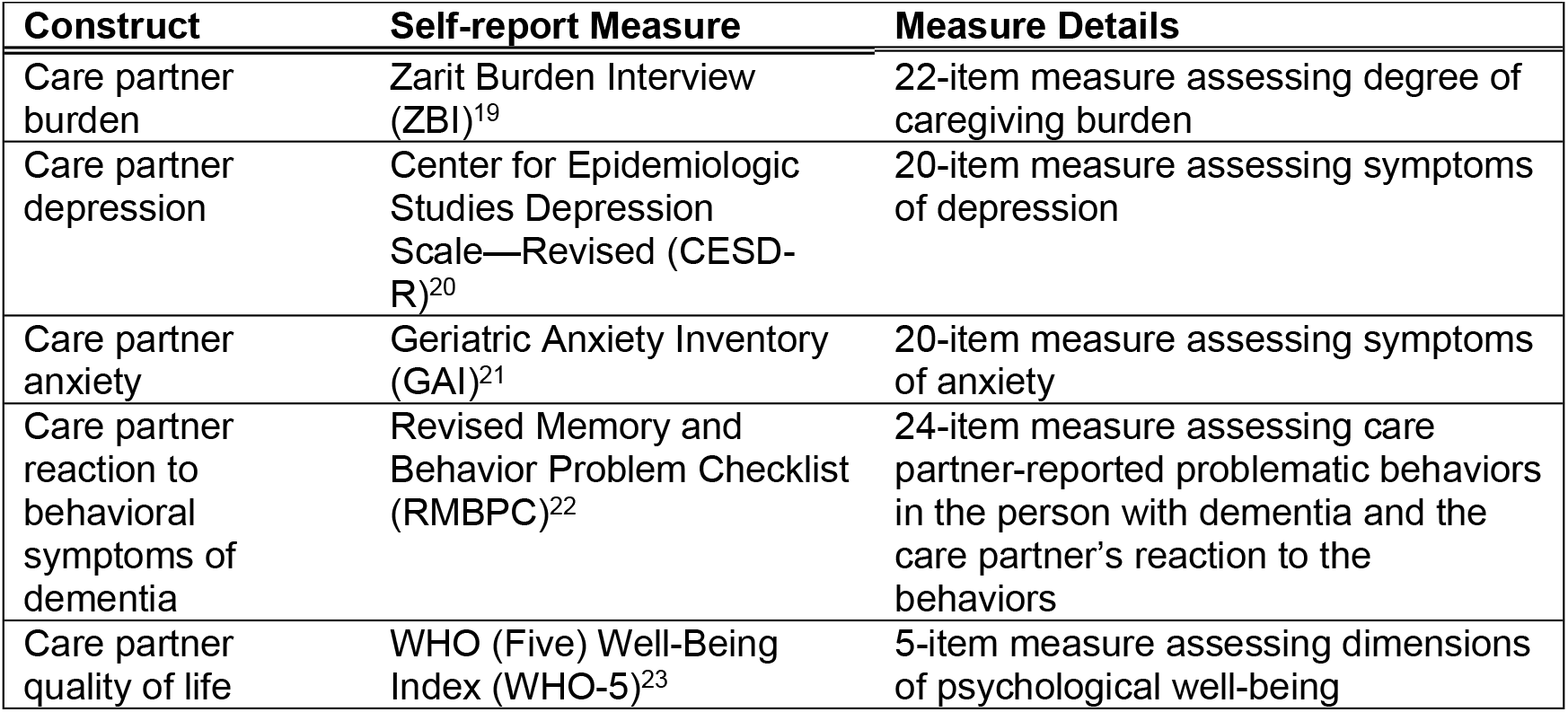
ICECaP Randomized Clinical Trial Primary Outcome Measures.

| Table 1. ICECaP Randomized Clinical Trial Primary Outcome Measures |  |  |
| --- | --- | --- |
| Construct | Self-report Measure | Measure Details |
| Care partner burden | Zarit Burden Interview (ZBI)<sup>19</sup> | 22-item measure assessing degree of caregiving burden |
| Care partner depression | Center for Epidemiologic Studies Depression Scale—Revised (CESD-R)<sup>20</sup> | 20-item measure assessing symptoms of depression |
| Care partner anxiety | Geriatric Anxiety Inventory (GAI)<sup>21</sup> | 20-item measure assessing symptoms of anxiety |
| Care partner reaction to behavioral symptoms of dementia | Revised Memory and Behavior Problem Checklist (RMBPC)<sup>22</sup> | 24-item measure assessing care partner-reported problematic behaviors in the person with dementia and the care partner's reaction to the behaviors |
| Care partner quality of life | WHO (Five) Well-Being Index (WHO-5)<sup>23</sup> | 5-item measure assessing dimensions of psychological well-being |

#### Intervention

After baseline questionnaires were completed, a trained dementia care coordinator (referred to going forward as care coordinator) called or e-mailed care partners in the ICECap group to schedule the initial contact. Care coordinators contacted the care partner at least once per month via email, telehealth phone/video, or an in-person meeting for at least 15 minutes. During these contacts, care coordinators provided supportive counseling and related services based on the needs of the care partner (e.g., behavioral management, safety strategies, case management, healthcare referrals, psychoeducation). Care partners were encouraged to contact care coordinators as needed. After the initial contact, care coordinators were also required to attend regular follow-up appointments in the Memory and Aging Care Clinic with the care partner and their associated person with dementia. These follow-up appointments typically occur every 6 to 12 months.

#### Control Group

Care partners in the control group received standard care as provided by the MACC (e.g., follow-up appointments every 6 to 12 months). They filled out the same questionnaires at baseline and 12 months as the care partners in the ICECaP intervention group.

### Statistical Analysis

#### Data summarization

Descriptive categorical data were summarized by frequencies (n) and relative frequencies (%), and descriptive continuous scaled data were summarized by the either the mean and standard deviation (SD) or the median and interquartile range of the distribution.

#### Baseline analysis

Although randomization imbalance with respect to the baseline demographic and caregiving characteristics of the ICECaP and control groups would expect to be rare, sensitivity analyses were conducted to assess for such imbalances. Statistical comparisons of distribution means were conducted via the Welch two-sample t-test, while statistical comparisons of distribution medians were conducted by the Wilcoxon two-sample Rank Sum test. Statistical comparisons of the relative frequencies of each characteristic were conducted by the Pearson two-sample exact test.

#### Efficacy analyses

Pre- to post-intervention 12-month changes in care partner burden (ZBI total score), depression (CESD-R total score), anxiety (GAI total score), reaction to dementia symptoms (RMBPC Reaction score) and quality of life (WHO-5 score) were the focus of the ICECaP trial efficacy analyses. Each efficacy analysis was conducted using a two-step analytical approach. In step 1, a linear mixed ANOVA model was used to estimate under the relaxed heterogenous, versus homogenous equal variance assumption, the intervention-specific mean pre- to post-intervention 12-month efficacy outcome change. A linear mixed model-derived one-sample t-test was then used to test the null hypothesis that the mean pre- to post-intervention 12-month efficacy outcome change is equal to zero (versus the alternative: not equal to zero). A two-sided α=0.05 significance level was used. In step 2, a linear mixed ANCOVA model was used to test the null hypothesis that the mean pre- to post-intervention 12-month efficacy outcome change is equal for the two groups after accounting for between-group disparities in the relevant baseline care partner outcome, in baseline Katz Basic ADLs, and in baseline Lawton-Brody Complex ADLs (versus the alternative: the mean pre- to post-intervention 12-month adjusted efficacy outcome change is not equal). A linear model adjusted contrasts of means was used to test the null hypothesis, and a two-sided α=0.05 significance level was used.

## Results

### Baseline Results

There were no significant group differences on baseline demographic and caregiving characteristics between the ICECaP and control groups (ps > .05 across comparisons; see Table 2).

**Table 2.** Care Partner Demographic and Caregiving Characteristics.

| Table 2. Care Partner Demographic and Caregiving Characteristics |  |  |  |
| --- | --- | --- | --- |
|  | ICECaP<br>(n=61) | Controls<br>(n=69) | P-value |
| Years of age [M(SD)] | 64.8 (12.3) | 64.9 (11.6) | 0.968 |
| Gender (women) | 42 (68.9%) | 52 (75.4%) | 0.438 |
| Race/Ethnicity |  |  |  |
| Non-Hispanic White | 57 (93.4%) | 60 (87.0%) | 0.790 |
| Non-Hispanic Black/African American | 3 (4.9%) | 4 (5.8%) |  |
| Hispanic/Latino | 1 (1.6%) | 3 (4.3%) |  |
| Non-Hispanic American Indian | 0 (0.0%) | 1 (1.4%) |  |
| Highest level of education completed |  |  |  |
| High school diploma or GED | 5 (8.2%) | 5 (7.2%) | 0.810 |
| Some college | 10 (16.4%) | 8 (11.6%) |  |
| Associate's degree | 4 (6.6%) | 8 (11.6%) |  |
| Bachelor's degree | 27 (44.3%) | 25 (36.2%) |  |
| Master's degree | 11 (18.0%) | 15 (21.7%) |  |
| Doctoral degree | 4 (6.6%) | 6 (8.7%) |  |
| Employment status* |  |  |  |
| Retired | 24 (39.3%) | 39 (56.5%) | 0.051 |
| Full-time | 15 (24.6%) | 15 (21.7%) |  |
| Part-time | 9 (14.8%) | 2 (2.9%) |  |
| Unemployed | 2 (3.3%) | 3 (4.3%) |  |
| Monthly income |  |  |  |
| \$1–\$4,999 | 24 (39.3%) | 19 (27.5%) | 0.255 |
| \$5,000–\$9,999 | 13 (21.3%) | 18 (26.1%) | |
| \$10,000–\$14,999 | 4 (6.6%) | 5 (7.2%) | |
| \$15,000–\$24,999 | 4 (6.6%) | 11 (15.9%) | |
| Relationship to PWD |  |  |  |
| Spouse/Partner | 44 (72.1%) | 49 (71.0%) | 1.000 |
| Adult Child | 14 (23.0%) | 16 (23.2%) |  |
| Co-dwelling with PWD | 47 (77.0%) | 54 (78.3%) | 0.688 |
| CP hours/month supporting basic ADLs for PWD [MD [IQR]] | 3.8 [0, 30.0] | 0 [0, 48.7] | 0.976 |
| CP hours/month supporting complex ADLs for PWD [MD [IQR]] | 30.0 [7.1, 9.0] | 60.0 [19.0, 120.0] | 0.067 |
| Katz Index of Independence in (Basic) ADLs for PWD [M(SD)] | 7.3 (0.9) | 7.6 (1.1) | 0.109 |

| **Table 2. Care Partner Demographic and Caregiving Characteristics** |  |  |  |
| --- | --- | --- | --- |
|  | **ICECaP**<br>**(n=61)** | **Controls**<br>**(n=69)** | **P-value** |
| Lawton-Brody Instrumental (Complex) ADL Scale for PWD [M(SD)] | 3.9 (2.2) | 3.7 (2.1) | 0.659 |
| Neuropsychiatric Inventory Questionnaire- PWD Severity Score [M(SD)]<sup>17</sup> | 6.1 (5.1) | 5.7 (4.7) | 0.630 |
| *Note.* Data presented as *n* (% of group) unless otherwise noted. ADLs = activities of daily living; CP = care partner; M(SD) = Mean (Standard Deviation); MD [IQR] = Median [Interquartile Range] PWD = person with dementia. |  |  |  |

There were no significant differences in baseline scores on any primary outcome measures between groups (see Table 3). Within both groups at baseline, caregiver burden fell in the mild to moderate range; mean depression scores and anxiety scores fell below the threshold for at-risk clinical depression and anxiety, respectively; mean reaction to behavioral symptoms of dementia fell between “a little” to “moderately” bothered by the person with dementia’s symptoms. There is no established cut-off for the WHO-5 quality of life score.

**Table 3.** ICECaP Intervention Primary 12-Month Efficacy Results.

| Table 3. ICECaP Intervention Primary 12-Month Efficacy Results |  |  |  |  |  |  |  |  |  |
| --- | --- | --- | --- | --- | --- | --- | --- | --- | --- |
|  | ICECaP (n=61) |  |  |  | Controls (n=69) |  |  |  | ANCOVA Models |
| Construct | Baseline Mean (SD) | 12-Month Mean (SD) | Mean $\Delta$ [95% CI] | P-value<sup>1</sup> | Baseline Mean (SD) | 12-Month Mean (SD) | Mean $\Delta$ [95% CI] | P-value<sup>2</sup> | P-value<sup>3</sup> |
| Burden | 17.30 (7.84) | 18.38 (7.82) | 1.25 [-0.56, 3.06] | 0.173 | 17.74 (7.96) | 18.74 (7.58) | 1.00 [-0.44, 2.44] | 0.170 | 0.955 |
| Depression | 9.11 (11.06) | 10.43 (10.83) | 1.33 [-1.55, 4.21] | 0.358 | 11.22 (11.15) | 13.14 (13.68) | 1.93 [-0.10, 3.96] | 0.062 | 0.563 |
| Anxiety | 4.46 (4.70) | 4.31 (4.61) | -0.15 [-1.28, 0.99] | 0.796 | 4.62 (4.81) | 5.01 (4.64) | 0.39 [-0.31, 1.10] | 0.272 | 0.396 |
| Reaction to dementia sx | 1.31 (0.69) | 1.25 (0.71) | -0.05 [-0.26, 0.16] | 0.621 | 1.34 (0.65) | 1.40 (0.65) | 0.06 [-0.09, 0.21] | 0.441 | 0.204 |
| Quality of life | 14.21 (5.13) | 13.95 (5.31) | -0.40 [-1.62, 0.82] | 0.515 | 13.77 (4.71) | 13.41 (5.20) | -0.38 [-1.41, 0.65] | 0.468 | 0.954 |
| Constructs were measured using the following questionnaires: Burden = ZBI-SF; Depression = CESD-R; Anxiety = GAI; Reaction to Dementia sx (Symptoms) = RMBPC Reaction score; Quality of Life = WHO-5<br>P-values<sup>1-2</sup> were derived from linear mixed model one-sample t-test for testing the null hypothesis that the mean change ($\Delta$) from baseline to 12-months is equal to zero. P-values<sup>3</sup> were derived from the linear mixed ANCOVA model two-sample test for comparing the mean change from baseline to 12-months ($\Delta$) between ICECaP and control groups after adjusting for baseline Katz Index of Independence in (Basic) ADLs and Lawton-Brody Instrumental (Complex) ADL Scale for PWD.<br>Analysis was by originally assigned groups. |  |  |  |  |  |  |  |  |  |

### Longitudinal Results

There were no significant differences in baseline to 12-month change on primary outcome measures between ICECaP and controls (see Table 3). Results were consistent when the Katz Basic ADLs and Lawton-Brody Complex ADLs were included as covariates in analyses.

### Harms

There were no harms associated with the intervention. Inherent harms of providing care for a person with dementia include possible depressive episodes.

## Discussion

This paper presents the preliminary, 12-month efficacy results of a pilot RCT of the ICECaP intervention for care partners of persons with dementia. On average, the 12-month intervention did not appear to significantly change care partners’ self-reported levels of burden, depression, anxiety, quality of life, or reaction to behavioral symptoms in the person with dementia. There were no significant differences between the ICECaP intervention group and the control group in key outcomes at baseline, 12-months, or in change from baseline to 12-months.

There are several possible explanations why there were no significant mental health, burden, or quality of life benefits detected in this 12-month RCT of ICECaP. First, this cohort of dementia care partners did not indicate experiencing elevated mental health distress, severe caregiving burden, or poor quality of life at baseline according to self-report measures. This is consistent with the relatively high socioeconomic status of this sample, who are majority non-Hispanic White and well educated, rendering them less vulnerable to adverse impacts of caregiving. Therefore, a floor effect may be present in which care partners’ self-report measure scores did not have ample room to allow for change in a positive (i.e., improved) direction.

Additionally, the relatively high socioeconomic status of the current sample limits generalizability of these findings to caregivers from more diverse socioeconomic backgrounds. In sum, future efficacy testing of ICECaP should have more restrictive inclusion criteria to include only care partners who have elevated levels of the target factor (e.g., burden, depression, and/or anxiety) and greater consideration of diverse socioeconomic statuses.

Additionally, no differential group-by-time effects may have been detected in this RCT due to the nature of the control group in this study. Specifically, all care partners included in both the intervention and control group received follow-up care in the UVA Memory and Aging Care Clinic (MACC). Follow-up care in MACC typically involves a one-hour appointment for the person with dementia and their care partner(s) one to two times per year with a multidisciplinary team of neuropsychologists, neuropsychology postdoctoral fellows, a nurse practitioner, a pharmacist, an occupational therapist, and a speech-language pathologist, among other specialties. While these appointments are scheduled for the person with dementia, the accompanying care partner receives information about pharmacological and non-pharmacological behavioral management strategies; long-term care planning support; dementia psychoeducation; psychotherapy and support group options for care partners; information regarding adult day, continuing care, and respite facilities; and other local resources. Further, care partners are often provided emotional support, validation, and encouragement; at times, the care partner and the person with dementia are separated during the appointment so a care partner can receive one-on-one validation and support (e.g., while the person with dementia is completing brief testing to inform treatment recommendations). Although once to twice yearly support in the multidisciplinary clinic is a lower frequency and intensity of support relative to the monthly support care partners receive in ICECaP, it is possible that there is too much overlap in support that was provided to both the ICECaP group and the control group to detect a signal. In the future, it may be more appropriate to restrict the control group to care partners in the community who are on the waitlist for specialty care services.

Another possible reason for the lack of differential changes on outcome measures between the ICECaP and control groups over time could be the variability in “dosage” of the intervention in the ICECaP group. As Thompson et al. (manuscript currently under review) reported, the number of contacts between dementia care coordinators and care partners in the ICECaP group was significantly variable. Care partners and care coordinators had an average of 2.2 contacts per month, averaged across 12-months within the ICECaP group; however, the number of contacts in a month ranged from 0 to 15 contacts. Further, while all care partners had a least one contact with a care coordinator during at least 11 of the 12 months of ICECaP, 25.6% of care partners had one month with 0 contacts with the care coordinator. In sum, it is possible that the intervention is efficacious for improving mental health, burden, and quality of life for those who are highly engaged in the program (e.g., > monthly contact with a care coordinator; care partner utilizes resources effectively by following through on care coordinator recommendations) but that engagement was too variable within the intervention group to detect improvement on outcome measures relative to the control group. Future analyses will focus on the extent to which engagement metrics impact efficacy of the intervention.

While the ICECaP pilot RCT did not demonstrate significant impacts of ICECaP on care partner mental health and caregiving burden, secondary analyses have revealed promising effects on other aspects of caregiver wellbeing (manuscript in preparation). Specifically, ICECaP appears to improve care partners’ preparedness for caregiving, which has downstream correlates with improvement in mental health measures. Collectively, the results from this manuscript and the forthcoming manuscript on ICECaP primary and secondary outcomes will provide important considerations for further refinement and testing of the ICECaP intervention.

## Data Availability

Data will be shared publicly upon completion of the full trial (approximately 2026). Limited datasets may be available upon request to the University of Virginia (contact Carol Manning).

## Acknowledgements

None.

